# Blood Draw Site and Analytic Device Influence Hemoglobin Measurements

**DOI:** 10.1101/2022.04.09.22273541

**Authors:** David W. Killilea, Frans A. Kuypers, Sandra K. Larkin, Kathleen Schultz

**Affiliations:** Office of Research, University of California, San Francisco, CA, USA; Division of Hematology, Department of Pediatrics, University of California, San Francisco, CA, USA

**Keywords:** anemia, hemoglobin, point-of-care instrument, blood collection tube

## Abstract

Anemia is a continuing global public health concern and a priority for international action. The prevalence of anemia is estimated from the hemoglobin (Hb) levels within target populations, yet the procedures for measuring Hb are not standardized and different approaches may result in discrepancies. Several analytical variables have been proposed to influence Hb measurements, but it is difficult to understand the impact on specific variables from large population or field studies. Therefore, we designed a highly controlled protocol that minimized most technical parameters to specifically investigate the impact of blood draw site and analytic device on Hb measurements. A diverse cohort of sixty healthy adults each provided a sequential capillary and venous blood sample that were measured for Hb using an automated hematology analyzer (ADVIA-2120) and two point-of-care devices (HemoCue 201+ and HemoCue 301). Comparing blood draw sites, the mean Hb content was 0.32-0.47 g/dL (2-4%) higher in capillary compared to venous blood from the same donors. Comparing different Hb measuring instruments, the mean Hb content was 0.19-0.46 g/dL (1-4%) higher measured with HemoCue devices compared to ADVIA-2120 in both capillary and venous blood from the same donors. The maximum variance in measurement was also higher with HemoCue devices using blood from venous (5-6% CV) and capillary (21-25% CV) sites compared to ADVIA-2120 (0.6-2% CV). Other variables including blood collection tube manufacturer did not affect mean Hb content. These results demonstrate that even when most technical variables are minimized, the blood draw site and the analytical device can have a small but statistically significant effect on the mean and dispersion of Hb measurements. Even in this study, the few participants identified as mildly anemic using venous blood measured by ADVIA-2120 would not have been classified as anemic using their capillary blood samples or point-of-care analyzers. Thus, caution is warranted when comparing Hb values between studies having differences in blood draw site and Hb measuring device. Future anemia testing should maintain consistency in these analytical variables.

## INTRODUCTION

Despite decades of international action, amenia is still a major health concern globally. The World Health Organization (WHO) estimates the prevalence of anemia at 20% worldwide, 30% for women aged 15-49 years, 40% for children aged 6-59 months overall, and 60% for children aged 6-59 months from low and middle-income countries (LMIC) [WHO 2001; Culleton 2006; World Health Organization 2008; Stevens 2013; Kassebaum 2016]. Anemia can lead to significant morbidities, including fatigue, reduced cognitive function, behavioral disturbances, and poor pregnancy outcomes [Viteri 1994; Silverberg 2001; Balarajan 2011]. Given the substantial impact on health and quality of life, many public health organizations regularly assess anemia levels in their target populations. These surveys depend on the accurate measurement of Hb to define the level of anemia, which is used to calculate the proportional amount of resources to support community health in these populations [WHO 2011]. Thus, it is critical to determine Hb levels with high precision and accuracy to determine baseline anemia levels and to subsequently evaluate the effect of intervention programs. Given the importance placed on Hb measurement, it is perhaps surprising that the collection and processing of blood for Hb measurement is not a standardized procedure, varying in a number of analytical variables that include how blood samples are drawn and processed, and what method is used for Hb measurement. This has led to concerns about the comparison of Hb values between different studies and best practices for future studies [Sari 2001; Cable 2012; Karakochuk 2015; Karakochuk 2019; Neufeld 2019; Whitehead 2019; Larson 2021].

Of the many analytical variables relevant to the determination of Hb levels, the two that have arguably seen the most change in recent years involves (1) the site of the blood draw and (2) the type of device used for Hb assessment. For blood draw site, the gold standard for assessing Hb levels is using venous blood obtained by standard phlebotomy [Neufeld 2019]. Phlebotomy utilizes a standard set of procedures and yields a substantial blood volume to complete the measurement of Hb and any other concomitant testing. However, phlebotomy has some disadvantages, including the need for trained technicians, specific sterile supplies, and potential adverse events after venipuncture. The most common alternative to phlebotomy is blood draw by fingerstick [Whitehead 2017]. Fingerstick sampling is a simple procedure that uses commercial lancet devices which are low cost, easy to operate, and have minimal adverse events after fingerstick. The main disadvantage to a fingerstick was the typical yield of a few drops of blood, which historically was insufficient for measurement. However, technological development has resulted in much lower blood volume requirements, so fingerstick sampling has become increasingly common for assessing anemia. The United States Agency for International Development, Centers for Disease Control and Prevention, and many other agencies routinely use fingerstick methods for blood sampling [Sharman 2000; ICF 2012; Pullum 2017].

Another changing preference in the measurement of Hb is the choice of analytical device. The gold standard for measuring Hb levels is the use of an automated hematology analyzer or similar instrument [Neufeld 2019]. These analyzers are robust, have excellent accuracy and reproducibility, and often measure other hematological parameters that provide a more complete clinical profile of the blood samples. However, automated hematology analyzers have some disadvantages, including high costs to purchase and maintain, need for technically proficient operators, and poor portability such that the use in field settings is difficult. In response to these challenges, handheld devices were developed nearly thirty years ago that could provide point-of-care (POC) testing for Hb concentrations using small amounts of whole blood [Whitehead 2019]. Extensive testing of the POC devices compared to automated hematology analyzers demonstrated that the POC devices had good accuracy and reproducibility for Hb, so they became preferred analytical devices for Hb measurements in large population studies and/or in field settings. Even the American Red Cross and other domestic clinical programs routinely use POC devices for anemia assessment [Osborn 2019; Whitehead 2019]. Two of the most successful devices are the HemoCue 201+ and HemoCue 301 (HemoCue America), which are still in common use today [Whitehead 2017]. A single drop of blood is sufficient for measuring Hb on these devices, which are easy to operate and do not require extensive technical training. The combination of fingerstick sampling and POC devices for Hb measurements creates significant convenience and cost-savings for anemia assessment in modern public health surveys.

The shift to fingerstick sampling and use of POC devices has not been without criticism. Fingerstick sampling draws capillary, not venous, blood, and studies have shown that the levels of blood components and biomarkers in capillary blood may differ from the levels in the venous circulation [Kaplan 1959; Falch 1981; Kupke 1981]. Many, although not all, of these studies have reported that Hb levels from capillary blood are elevated compared to venous blood [Neufeld 2019; Whitehead 2019]. Additionally, there are some reports showing discrepancies and higher variability when Hb was measured using POC devices compared to traditional automated hematology analyzers [Neufeld 2019; Whitehead 2019]. Yet in most of these cases, multiple technical variables can be identified which might impact the determination of Hb – including difference in phlebotomists, supply chain, or clinical settings – making it difficult to assess which parameters were most responsible for the discrepancies in Hb measurement. We aimed to focus on the impact of blood draw site and measuring device on Hb measurement in an analysis without complications from technical and human factors, or from the clinical environment. Therefore, we collected blood samples from a healthy adult cohort in a highly controlled study while minimizing extraneous variables. Our results show how blood draw site and measurement device can affect reported Hb values, and why caution is justified when comparing Hb values between studies using different Hb assessment protocols.

## RESULTS

### Collection of blood samples

A cohort of 60 healthy adults was recruited from the local community to provide blood donations for the comparison of analytic variables that may influence Hb measurements. The participants were diverse in terms of age, gender, race/ethnicity, and other factors (**Supplemental Table 1 & Supplemental Figure 1**). Most technical variables were controlled, including use of the same phlebotomist, lab technician, blood draw procedure, and other parameters that commonly vary in other studies (**Table 1**). After sharing basic demographic and anthropomorphic data, each participant provided a capillary blood sample by fingerstick and then immediately provided venous blood sample by venipuncture. Blood samples were collected into a series of blood collection tubes (BCTs) from the same tube types and lot numbers, and all samples were processed in the nearby laboratory in under 60 minutes, with most processed within 10-20 minutes. Using a modified protocol for capillary blood collection, we were able to collect capillary blood samples from all 60 participants, with 52 providing over 1ml, which was enough volume for testing on the automated hematology analyzer and POC instruments. All procedural details were recorded for each participant, including venipuncture location, estimated volume, and time until Hb measurement.

**Table 1.**
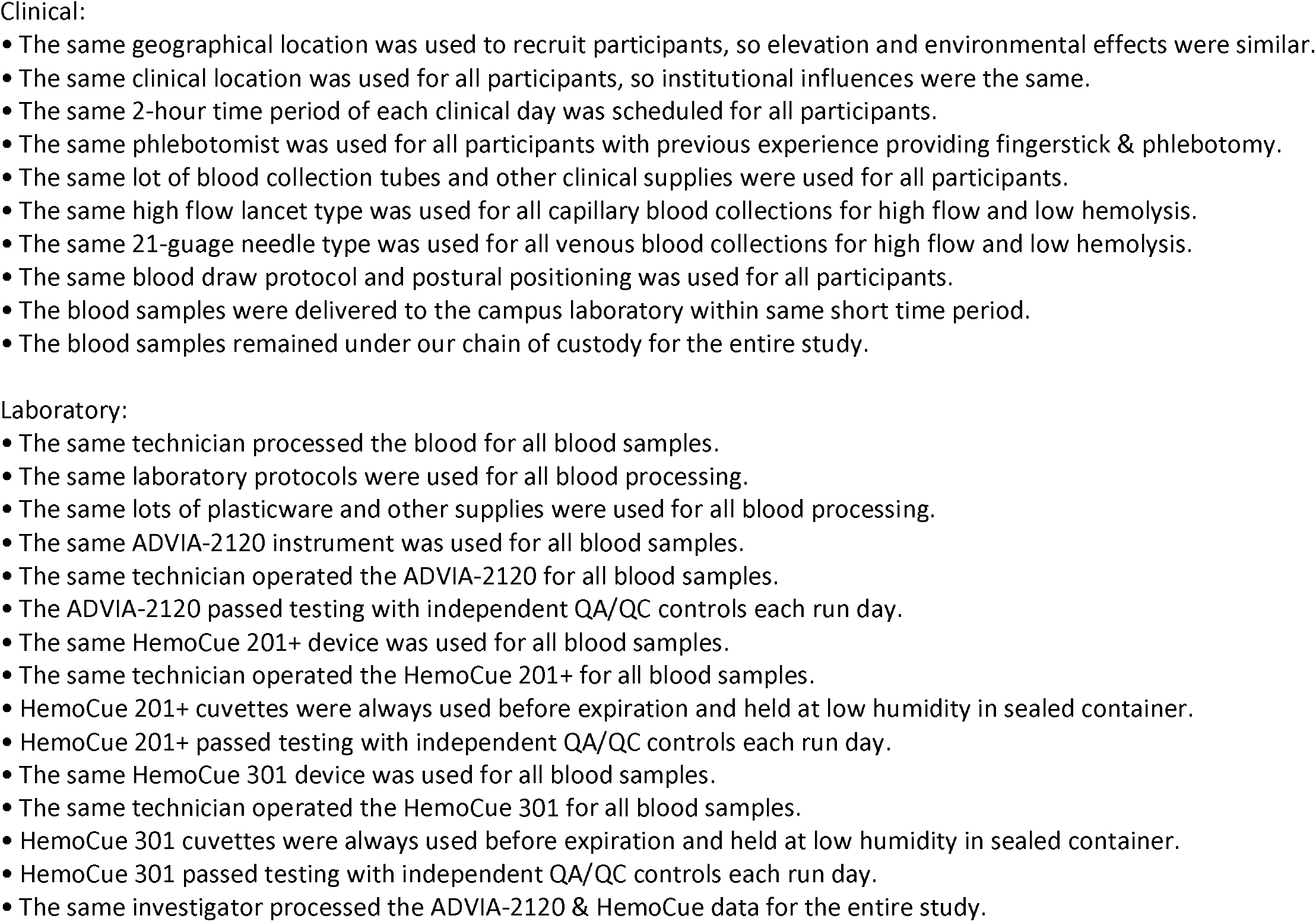
Analytical variables controlled in this study.

### Assessment of hemoglobin data

The gold standard for Hb assessment is the use of venous blood assessed using an automated hematology analyzer, so these data were evaluated first (**Figure 1**). The Hb levels in venous blood from all participants measured by ADVIA-2120 ranged between 10.8-17.0 g/dL, were normally distributed, and had a mean of 14.22 ± 1.28 g/dL (n=60) with a CV of 9.0%. There were no improbable values observed using the criteria of Hb >19.0 g/dL. When separated by sex of participants, the mean Hb for female participants (13.54 ± 1.07 g/dL) was lower than male (15.05 ± 1.01 g/dL), which was significantly different by two-tailed unpaired t test (p<0.0001). Using a common Hb threshold for anemia in US non-smoking adults of 11.7 g/dL [Tietz 2006], 3 participants would be classified as having mild anemia despite reporting being in good health. Although the anemic participants were all female, removing these values from the female group resulted in a mean Hb of 13.78 ± 0.78 g/dL with CV of 5.6%, which was still significantly different than the male-group mean Hb by two-tailed unpaired t test (p<0.0001). There was no significant difference in mean Hb content between participants based on age, race/ethnicity, and body mass index (BMI), data not shown.

**Figure 1.**
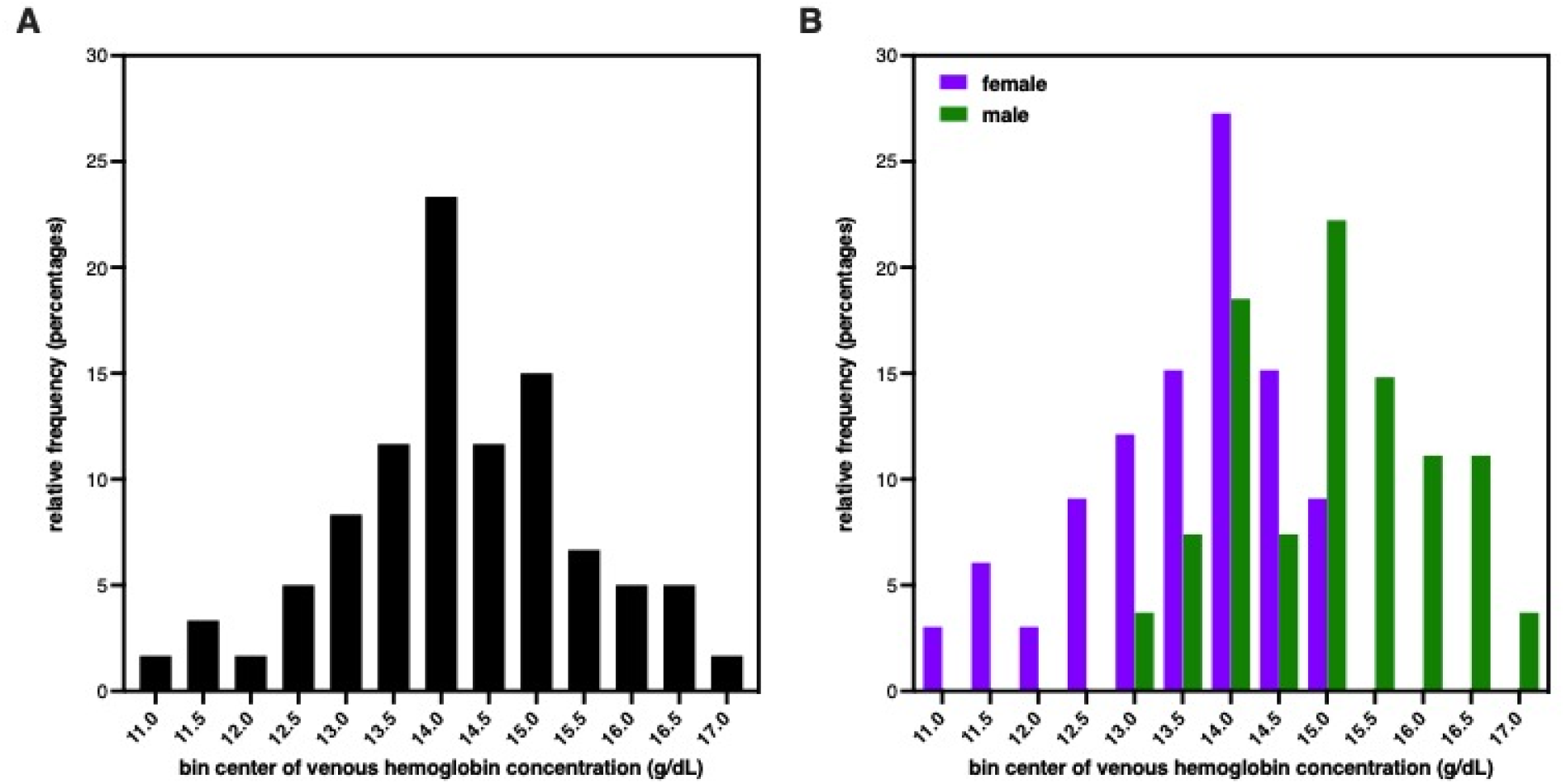
Distribution of hemoglobin concentration in study population. **(A):** Histogram of group Hb values determined from venous blood using ADVIA-2120. The mean Hb for all participants (n=60) was 14.22 ± 1.28 g/dL. **(B):** Histogram of Hb values from females (purple) and males (green) determined from venous blood using ADVIA-2120. The mean Hb for female participants (13.54 ± 1.07 g/dL, n=33) was significantly lower than male (15.05 ± 1.01 g/dL, n=27), p<0.0001.

### Comparison of blood draw site

For comparisons of Hb values from capillary or venous blood, the ADVIA-2120 was used as the primary measurement device. All 60 participants provided venous blood samples, but only 52 participants had sufficient volume of capillary blood for these measurements. No improbable Hb values were identified with the ADVIA-2120 and the data passed normality testing, so parametric statistics were used. The mean Hb for the blood samples for individuals with both venous and capillary assessment was 14.58 ± 1.43 g/dL Hb for capillary and 14.25 ± 1.31 g/dL Hb for venous (n=52), revealing that capillary Hb values were elevated by 0.33 g/dL in a sequential blood draw from the same participants (**Table 2 & Figure 2A**). This difference was statistically significant (p=0.0065) using a two-tailed paired t-test. Bland-Altman analysis revealed a similar bias of 0.34 ± 0.85 g/dL for capillary Hb values compared to venous Hb values.

**Table 2.**
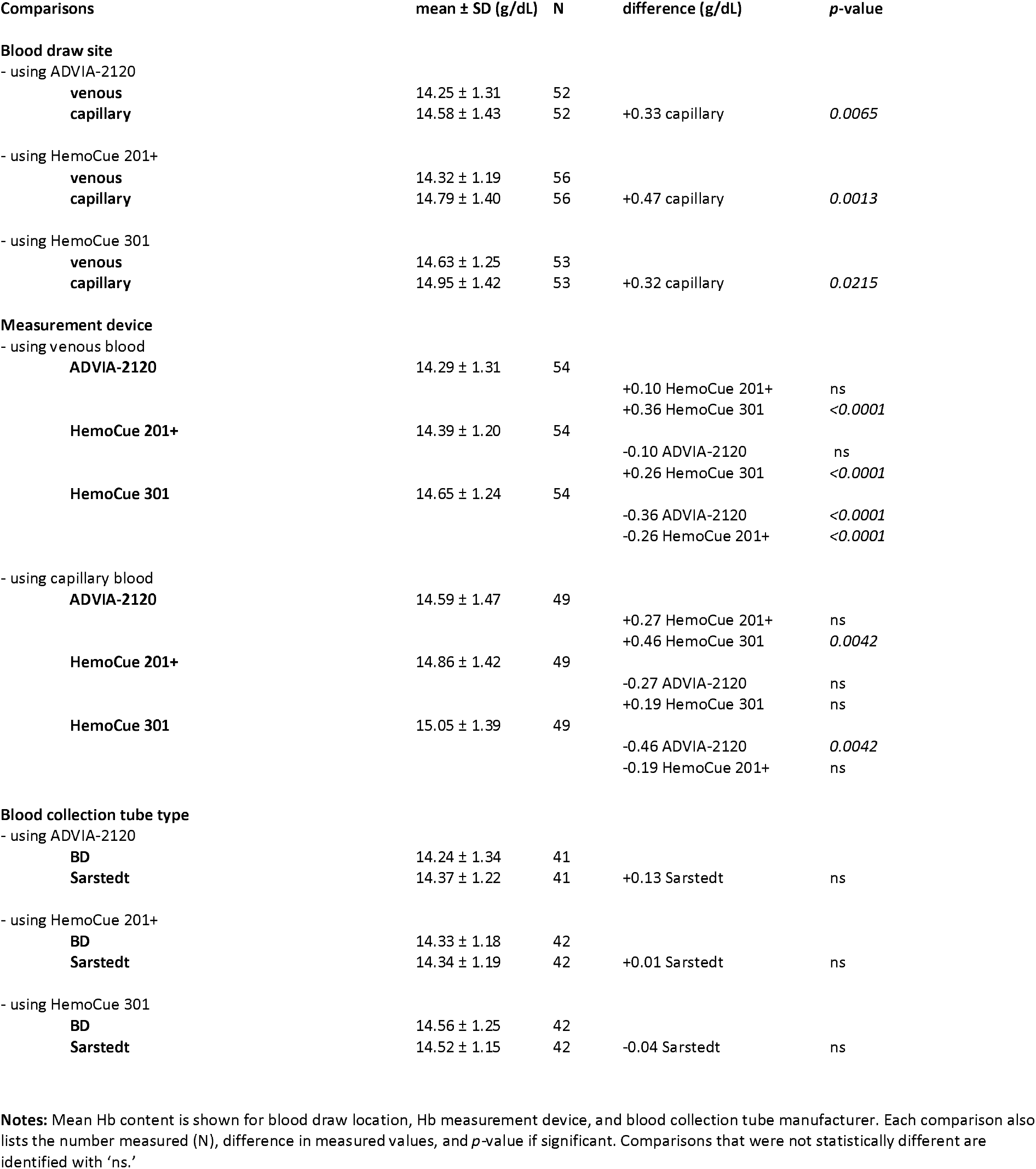
Hemoglobin measurements differ by blood draw site and measurement device, but not blood collection tube manufacturer.

| Comparisons | mean ± SD (g/dL) | N | difference (g/dL) | *p*-value |
| --- | --- | --- | --- | --- |
| **Blood draw site** |  |  |  |  |
| - using ADVIA-2120 |  |  |  |  |
| venous | 14.25 ± 1.31 | 52 | | |
| capillary | 14.58 ± 1.43 | 52 | +0.33 capillary | 0.0065 |
| - using HemoCue 201+ |  |  |  |  |
| venous | 14.32 ± 1.19 | 56 | | |
| capillary | 14.79 ± 1.40 | 56 | +0.47 capillary | 0.0013 |
| - using HemoCue 301 |  |  |  |  |
| venous | 14.63 ± 1.25 | 53 | | |
| capillary | 14.95 ± 1.42 | 53 | +0.32 capillary | 0.0215 |
| **Measurement device** |  |  |  |  |
| - using venous blood |  |  |  |  |
| ADVIA-2120 | 14.29 ± 1.31 | 54 | | |
|  |  |  | +0.10 HemoCue 201+ | ns |
|  |  |  | +0.36 HemoCue 301 | <0.0001 |
| HemoCue 201+ | 14.39 ± 1.20 | 54 | | |
|  |  |  | -0.10 ADVIA-2120 | ns |
|  |  |  | +0.26 HemoCue 301 | <0.0001 |
| HemoCue 301 | 14.65 ± 1.24 | 54 | | |
|  |  |  | -0.36 ADVIA-2120 | <0.0001 |
|  |  |  | -0.26 HemoCue 201+ | <0.0001 |
| - using capillary blood |  |  |  |  |
| ADVIA-2120 | 14.59 ± 1.47 | 49 | | |
|  |  |  | +0.27 HemoCue 201+ | ns |
|  |  |  | +0.46 HemoCue 301 | 0.0042 |
| HemoCue 201+ | 14.86 ± 1.42 | 49 | | |
|  |  |  | -0.27 ADVIA-2120 | ns |
|  |  |  | +0.19 HemoCue 301 | ns |
| HemoCue 301 | 15.05 ± 1.39 | 49 | | |
|  |  |  | -0.46 ADVIA-2120 | 0.0042 |
|  |  |  | -0.19 HemoCue 201+ | ns |
| **Blood collection tube type** |  |  |  |  |
| - using ADVIA-2120 |  |  |  |  |
| BD | 14.24 ± 1.34 | 41 | | |
| Sarstedt | 14.37 ± 1.22 | 41 | +0.13 Sarstedt | ns |
| - using HemoCue 201+ |  |  |  |  |
| BD | 14.33 ± 1.18 | 42 | | |
| Sarstedt | 14.34 ± 1.19 | 42 | +0.01 Sarstedt | ns |
| - using HemoCue 301 |  |  |  |  |
| BD | 14.56 ± 1.25 | 42 | | |
| Sarstedt | 14.52 ± 1.15 | 42 | -0.04 Sarstedt | ns |

**Notes:** Mean Hb content is shown for blood draw location, Hb measurement device, and blood collection tube manufacturer. Each comparison also lists the number measured (N), difference in measured values, and *p*-value if significant. Comparisons that were not statistically different are identified with 'ns.'

**Figure 2.**
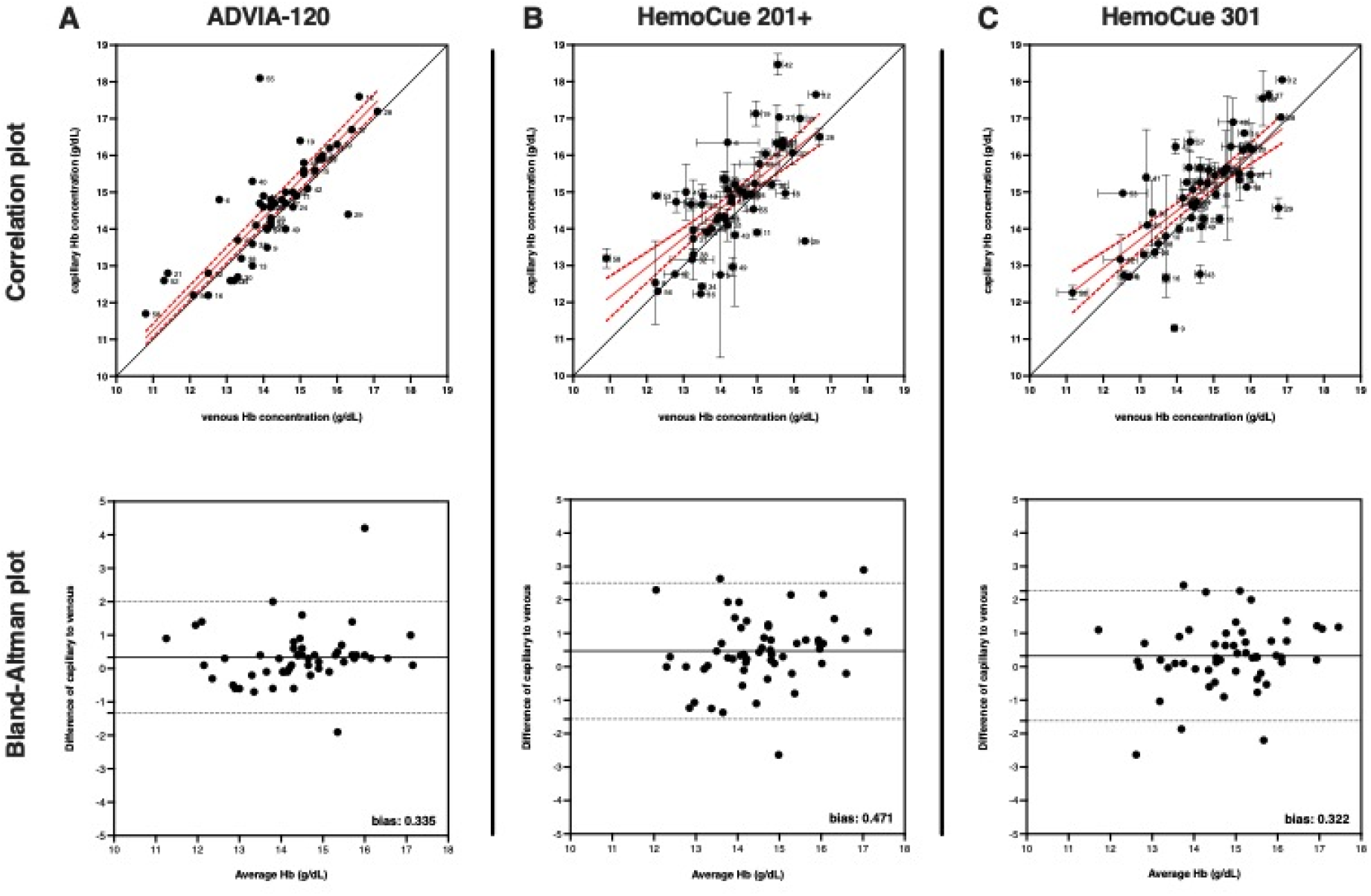
Hemoglobin concentration is elevated in capillary compared to venous blood. Correlation and Bland-Altman plots for Hb values determined in capillary and venous blood from the same participants measured by (**A**) ADVIA-2120, (**B**) HemoCue 201+, and (**C**) HemoCue 301. The correlation plots show the capillary and venous Hb values (mean ± SEM) proximity to line of concordance (solid black line), as well as linear regression (solid red line) ± 95% confidence interval (red dotted line) for the group. Each circle represents a single participant, with participant ID number to the immediate right. The Bland-Altman plot shows a bias for capillary over venous values as (**A**) 0.335 ± 0.850 g/dL for ADVIA-2120, (**B**) 0.471 ± 1.039 g/dL for HemoCue 201+, and (**C**) 0.322 ± 0.990 g/dL for HemoCue 301.

Differences in Hb measurement between capillary or venous blood were also tested using the POC devices. The HemoCue 201+ was used to measure Hb content in both capillary or venous blood. Fifty-seven participants were able to provide sufficient volume for both capillary and venous blood for these measurements. One sample was identified as an improbable value (Hb >19 g/dL) in the dataset, so 56 participants values were used for this comparison. These Hb data passed normality testing so parametric statistics were used. The mean Hb for the participants was 14.79 ± 1.40 g/dL Hb for capillary and 14.32 ± 1.19 g/dL Hb for venous, indicating that capillary Hb values were elevated by 0.47 g/dL in a sequential blood draw from the same participants (**Table 2 & Figure 2B**). This difference was statistically significant (p=0.0024) using a two-tailed paired t-test. Bland-Altman analysis revealed a similar bias of 0.47 ± 1.04 g/dL for capillary Hb values compared to venous Hb values.

The HemoCue 301 was also used to measure Hb content in capillary or venous blood. 53 participants were able to provide sufficient volume for both capillary and venous blood for these measurements. No improbable values were identified, so all 53 participant values were used for these measurements. These Hb data passed normality testing so parametric statistics were used. The mean Hb for the participants was 14.95 ± 1.42 g/dL Hb for capillary and 14.63 ± 1.25 g/dL Hb for venous, indicating that capillary Hb values were elevated by 0.32 g/dL in a sequential blood draw from the same participants (**Table 2 & Figure 2C**). This difference was statistically significant (p=0.0345) using a two-tailed paired t-test. Bland-Altman analysis revealed a similar bias of 0.32 ± 0.99 g/dL for capillary Hb values compared to venous Hb values.

In summary, capillary Hb values were 2-4% greater than venous Hb values taken from sequential blood draw from the same participants, depending on which measurement device was used.

### Comparison of Hb measurement device

For comparisons of Hb values using different measuring instruments, both capillary and venous blood samples were tested on the ADVIA-2120, HemoCue 201+, and HemoCue 301. The 201+ and 301 models were selected since they are arguably the most commonly used POC devices in large population studies [Whitehead 2019]. Fifty-four participants were able to provide sufficient volume of venous blood for these measurements. The Hb data from each instrument passed normality testing so parametric statistics were used. The mean Hb values determined from the different instruments was 14.29 g/dL ± 1.31 g/dL Hb from ADVIA-2120, 14.39 g/dL ± 1.20 g/dL Hb from HemoCue 201+, and 14.65 g/dL ± 1.24 g/dL Hb from HemoCue 301 (**Table 2**). The difference between mean Hb values measured by ADVIA-2120 and HemoCue 201+ was not statistically significant (p=0.4072). The difference between mean Hb values measured by ADVIA-2120 and HemoCue 301 was 0.36 g/dL and was statistically significant (p<0.0001). The difference between mean Hb values measured by HemoCue 201+ and HemoCue 301 was 0.26 g/dL and was statistically significant (p<0.0001). Correlation and Bland-Altman analysis were used to compare each of the devices to one another to evaluate bias (**Figure 3, panel A-C**). Comparison of the Hb values from the ADVIA-2120 and HemoCue 201+ revealed a bias of 0.03 ± 0.78 g/dL for HemoCue 201+ over ADVIA-2120. Comparison of the Hb values from ADVIA-2120 and HemoCue 301 revealed a bias of 0.36 ± 0.56 g/dL for HemoCue 301 over ADVIA-2120. Comparison of the Hb values from HemoCue 201+ and HemoCue 301 revealed a bias of 0.26 ± 0.37 g/dL for HemoCue 301 over HemoCue 201+.

**Figure 3.**
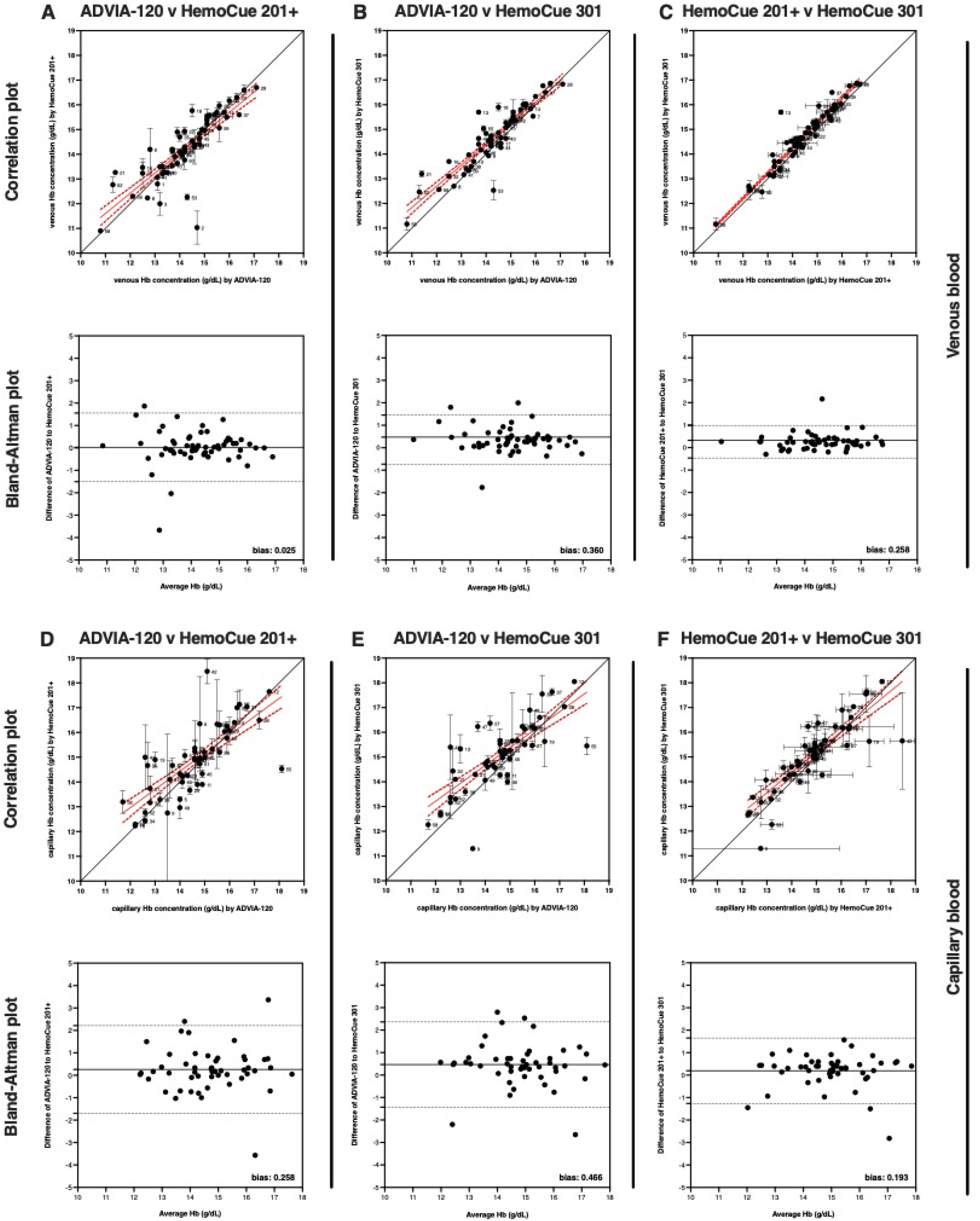
Hemoglobin concentration is elevated when measured by POC devices compared to an automated hematology analyzer. Correlation and Bland-Altman plots comparing venous Hb values measured by (**A**) ADVIA-2120 and HemoCue 201+, (**B**) ADVIA-2120 and HemoCue 301, and (**C**) HemoCue 201+ and HemoCue 301, and comparing capillary Hb values measured by (**D**) ADVIA-2120 and HemoCue 201+, (**E**) ADVIA-2120 and HemoCue 301, and (**F**) HemoCue 201+ and HemoCue 301. The correlation plots show the capillary and venous Hb values (mean ± SEM) proximity to line of concordance (solid black line), as well as linear regression (solid red line) ± 95% confidence interval (red dotted line) for the group. Each circle represents a single participant, with participant ID number to the immediate right. Using venous blood, the Bland-Altman plot shows a bias of (**A**) 0.025 ± 0.781 g/dL for HemoCue 201+ over ADVIA-2120, (**B**) 0.360 ± 0.558 g/dL for HemoCue 301 over ADVIA-2120, and (**C**) 0.258 ± 0.370 g/dL for HemoCue 301 over HemoCue 201+ measurements. Using capillary blood, the Bland-Altman plot shows a bias of (**D**) 0.258 ± 0.998 g/dL for HemoCue 201+ over ADVIA-2120, (**E**) 0.466 ± 0.968 g/dL for HemoCue 301 over ADVIA-2120, and (**F**) 0.193 ± 0.743 g/dL for HemoCue 301 over HemoCue 201+ measurements.

For comparisons of capillary Hb values from the 3 different measuring instruments, 49 participants were able to provide sufficient volume of capillary blood for these measurements. The Hb data passed normality testing so parametric statistics were used. The mean capillary Hb values determined from the 3 instruments was 14.59 g/dL ± 1.47 g/dL Hb from the ADVIA-2120, 14.86 g/dL ± 1.42 g/dL Hb from the HemoCue 201+, and 15.05 g/dL ± 1.39 g/dL Hb from the HemoCue 301 (**Table 2**). The difference between the mean Hb values measured by ADVIA-2120 and HemoCue 201+ was not statistically significant (p=0.1418). The difference between mean Hb values measured by ADVIA-2120 and HemoCue 301+ was 0.46 g/dL and was statistically significant (p=0.0042). The difference between mean Hb values measured by HemoCue 201+ and HemoCue 301+ was not statistically significant (p=0.1742). Correlation and Bland-Altman analysis were used to compare each of the devices to evaluate bias (**Figure 3, panel D-F**). Comparison of the Hb values from ADVIA-2120 and HemoCue 201+ revealed a bias of 0.26 ± 1.00 g/dL for HemoCue 201+ over ADVIA-2120. Comparison of the Hb values from ADVIA-2120 and HemoCue 301 revealed a bias of 0.47 ± 0.97 g/dL for HemoCue 301 over ADVIA-2120. Comparison of the Hb values from HemoCue 201+ and HemoCue 301 revealed a bias of 0.19 ± 0.74 g/dL for HemoCue 301 over HemoCue 201+.

In summary, Hb values from venous blood were greater when measured by HemoCue 301 compared to ADVIA-2120 or HemoCue 201+ when taken from the same blood draw from the same participants. Hb values from capillary blood were greater when measured by HemoCue 301 compared to ADVIA-2120 alone when taken from the same blood draw from the same participants. Using the results from venous blood as baseline, the observed difference of 0.36 g/dL represents a 2-3% increase in Hb values when using HemoCue 301 compared to ADVIA-2120. A similar increase in mean Hb (0.46 g/dL) is observed when measuring capillary blood with HemoCue 301 compared to ADVIA-2120. Additionally, the observed increase of 0.26 g/dL for venous blood represents a 1-2% increase in Hb values when using HemoCue 301 compared to HemoCue 201+, but this was not observed using capillary blood.

### Assessment of instrumentation variability

Comparison of Hb measurements with different devices also included an assessment of variability with replicate readings using the same blood samples. Improbable Hb values (<19 g/dL) were removed from the dataset. The remaining Hb data passed normality testing so parametric statistics were used. Repeated measurements of the same venous blood samples run during the study period using the ADVIA-2120 revealed a mean CV of 1.34 ± 0.78% (n=4) with a range between 0.55%-2.41%. Due to the larger volume requirements of the ADVIA-2120, only one capillary blood sample per participant was possible. Since the sample volume needed for the HemoCue 201+ and HemoCue 301 is much smaller, 3 replicates of the same capillary and venous blood samples were measured for most participants (**Figure 4**). For venous blood samples, repeated measures of blood samples using the HemoCue 201+ revealed a mean CV of 1.42 ± 1.35% (n=57) with a range between 0.39-6.04% and repeated measures of blood samples using the HemoCue 301 revealed a mean CV of 1.33 ± 1.12% (n=50) with a range between 0.36-5.52%CV. Capillary blood samples had a higher mean and maximum variability, as repeated measures of blood samples using the HemoCue 201+ revealed a mean CV of 4.00 ± 5.43% (n=54) with a range between 0.39-24.96% and repeated measures of blood samples using the HemoCue 301 revealed a mean CV of 4.01 ± 4.38% (n=48) with a range between 0.34-20.89%.

**Figure 4.**
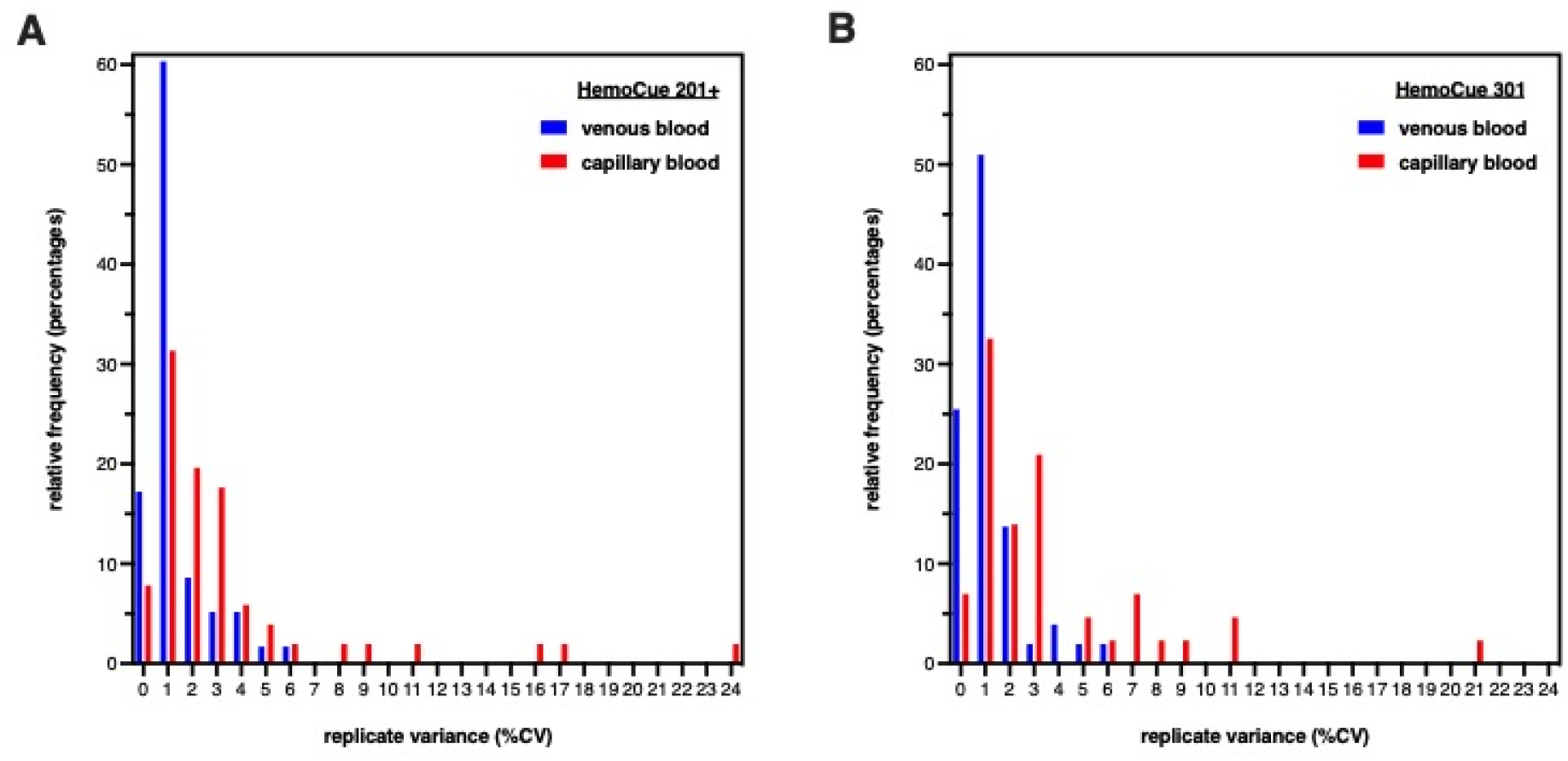
Variance in hemoglobin measurement replicates is greater from POC devices compared to automated hematology analyzers. Histograms of Hb values determined from replicate measurements of venous and capillary blood using (**A**) HemoCue 201+ and (**B**) HemoCue 301. For HemoCue 201+, the mean CV was 1% and the maximum CV was 6% (n=57) for venous samples (blue), whereas the mean CV was 4% and the maximum CV was 25% (n=54) for capillary samples (red). For HemoCue 301, the mean CV was 1% and the maximum CV was 6% (n=50) for venous samples (blue), whereas the mean CV was 4% and the maximum CV was 21% (n=48) for capillary samples, The reference values for mean CV of replicate Hb measurements using ADVIA-2120 is 0.6-2%.

### Comparison of BCT manufacturer

Becton, Dickinson and Company (BD) was selected as the manufacturer of BCTs used for this study because this vendor has the major market share in the US, but we were also able to test BCTs manufactured by Sarstedt, which has a major market share in European countries. Also, the anticoagulant type selected was ethylenediaminetetraacetic acid (EDTA) due to the routine use of this BCT format for whole blood analysis. Using venous blood samples, the Hb data passed normality testing so parametric statistics were used. For BD BCTs, the mean Hb values determined from the different instruments was 14.24 ± 1.34 g/dL Hb (n=41) from ADVIA-2120, 14.33 ± 1.18 g/dL Hb (n=42) from HemoCue 201+, and 14.56 ± 1.25 g/dL Hb (n=42) from HemoCue 301 (**Table 2**). For Sarstedt BCTs, the mean Hb values determined from the different instruments was 14.37 ± 1.22 g/dL Hb (n=41) from ADVIA-2120, 14.34 ± 1.19 g/dL Hb (n=42) from HemoCue 201+, and 14.52 ± 1.15 g/dL Hb (n=42) from HemoCue 301. These differences between BCT manufacturers ranged from 0.04 to 0.13 g/dL depending on measurement device, but none were statistically significant. Bland-Altman analysis revealed a small bias as well (**Supplemental Figure 2**). The limited volume of capillary samples precluded normality testing so non-parametric statistics were used. For the ADVIA-2120, no capillary blood samples were available for comparison due to the larger sample volume requirement of the instrument. For HemoCue 201+, only 5 participants provided enough capillary blood for these measurements. The mean Hb from BD BCTs was 13.73 ± 1.12 g/dL and the mean from Sarstedt BCTs was 13.80 ± 1.50 g/dL, which was not significantly different using a Wilcoxon test (p>0.9999). For HemoCue 301, only 4 participants provided enough capillary blood for these measurements. The mean Hb from BD BCTs was 13.98 ± 1.22 g/dL and the mean from Sarstedt BCTs was 13.43 ± 1.17 g/dL, which was also not significantly different using a Wilcoxon test (p=0.1250).

### Impact of blood draw site and measurement device on Hb values

The blood draw site and Hb measurement device both had a small but statistically significant effect on reported Hb values, so we evaluated how those variables might differentially affect Hb levels at the boundary of anemia. Although this study was not designed to evaluate anemic individuals, 3 participants had Hb values below 11.7 g/dL using the conventional venous blood measured on an automatic hematology analyzer and would therefore be classified as mildly anemic. Yet, using capillary blood, all 3 participants had Hb values at or above 11.7 g/dL and thus would not have been classified as anemic (**Table 3**). Additionally, the Hb values from venous blood measured by the HemoCue devices was also higher, resulting in 2 of the 3 participants also not being classified as anemic.

**Table 3.**
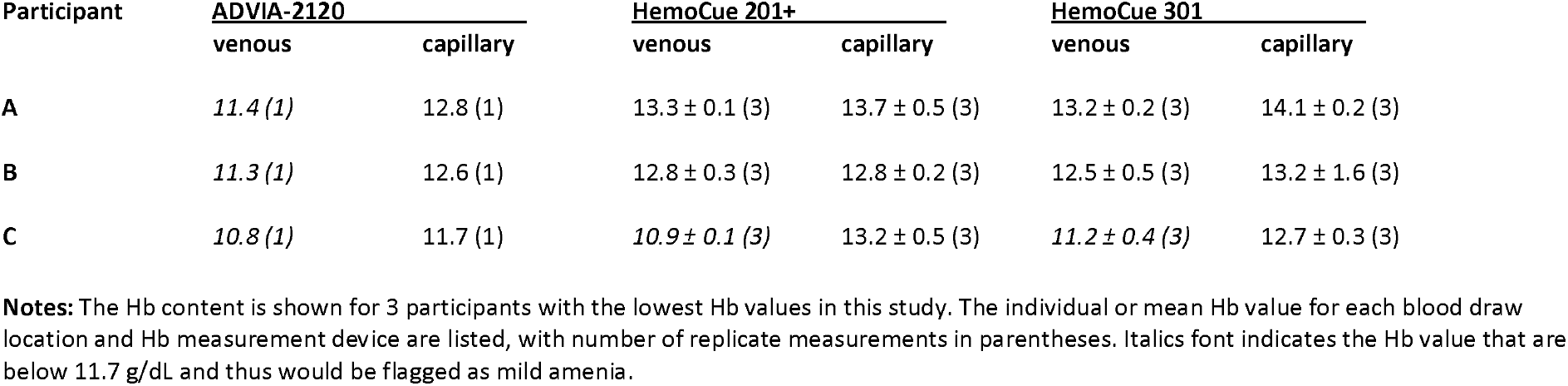
Hemoglobin measurements in select participants using values from different blood draw sites and measurement devices.

## DISCUSSION

This study was focused on how the blood draw site and analytic device can affect the assessment of anemia. Historically, Hb levels were measured in venous blood using automatic hematology analyzers or similar instruments, but newer POC devices have low blood volume requirements and portability, which allows for more widespread evaluation of Hb levels using capillary blood. Yet the current Hb thresholds for anemia are still based on venous blood read on automatic hematology analyzers, so it is important to quantify any bias inherent in capillary blood readings by POCs. A number of excellent studies have looked at this question, and the majority have noted small but significant differences in Hb values from these methods [Sari 2001; Cable 2012; Karakochuk 2015; Karakochuk 2019; Neufeld 2019; Whitehead 2019; Larson 2021]. However, those reports evaluated Hb data from large populations and/or field research with multiple complex analytical variables that are difficult to minimize, making it challenging to assign specific bias on any one parameter. Therefore, we used a more-controlled approach to identify the impact of different blood draw sites and analytic devices on Hb measurement.

The Hb data was analyzed from venous blood measured by the ADVIA-2120 automatic hematology analyzer as the gold standard. The full group mean was 14.22 ± 1.28 g/dL with a CV of 9.0%. When separated by sex, the mean Hb was 13.54 ± 1.07 g/dL for female participants and 15.05 ± 1.01 g/dL for male participants. These results were similar to other studies of anemia in the US population, including an analysis combining data from five National Health and Nutrition Examination Surveys (NHANES) from 2003 to 2012 (n=41,026) which found the overall mean Hb was 14.2 g/dL, with 13.4 g/dL for female and 14.9 g/dL for male participants [Li 2016]. Three female participants in this study were categorized as having mild anemia (Hb 11.0-11.8 mg/dL) despite reporting being in good health. Hb values in subgroups based on sex, with or without participants with anemia, were statistically different. Hb values in subgroups based on age, ethnicity, or BMI were not significantly different, but these group sizes were smaller and had low statistical power.

First, we determined how the blood draw site influenced the measurement of Hb, and found that the mean Hb content was 0.33 g/dL higher in capillary compared to venous blood from the same donors when measured by ADVIA-2120. Similar results of elevated capillary Hb were seen using HemoCue 201+ or HemoCue 301, so that capillary Hb levels were 2-4% higher than venous samples regardless of measurement device. Several previous studies have compared Hb levels from capillary and venous blood, and our findings are consistent with the majority of these showing elevated Hb levels in capillary samples [Chen 1992; Neufeld 2002; Cable 2012; Patel 2013; Chaudhary 2017; Neufeld 2019; Whitehead 2019; Larson 2021]. The reasons for the elevated capillary Hb values are not understood, but may include different flow characteristics of a colloidal suspension (blood) through vessels of disparate diameters, impact of interstitial fluid dynamics on capillary beds, and changes in vascular physiology due to sex or age [Neufeld 2019]. Without a clear understanding of the cause for divergent Hb values from venous and capillary sampling, it is difficult to set an adjustment factor or an approach to mitigate these differences. Larson and colleagues even concluded that “capillary and venous samples cannot be used interchangeably for the measurement of haemoglobin concentration and estimation of anaemia prevalence given the bias and imprecision when comparing the two. These findings call the global estimates of anaemia prevalence, which have been predominantly generated using capillary measurements, into doubt” [Larson 2021].

Next, we examined how the analytic device influenced the measurement of Hb, comparing venous and capillary blood from the same participants measured on the ADVIA-2120, HemoCue 201+, and HemoCue 301 instruments. Using venous blood, mean Hb content was 0.36 g/dL higher when measured by HemoCue 301 compared to ADVIA-2120, and 0.26 g/dL higher when measured by HemoCue 301 compared to HemoCue 201+. Using capillary blood, mean Hb content was 0.46 g/dL higher when measured by HemoCue 301 compared to ADVIA-2120. So regardless of blood draw location, the Hb values were 1-4% higher when measured by HemoCue 301 compared to the ADVIA-2120. Several previous studies have compared Hb measurements using automatic hematology analyzers to POC devices, and our findings are consistent with many showing elevated Hb levels in some HemoCue models under specific conditions [Paiva 2004; Karakochuk 2015; Chaudhary 2017; Boghani 2017; Hinnouho 2017; Whitehead 2017; Karakochuk 2019; Whitehead 2019], while other studies did not note significant differences [Larson 2021]. The reasons for the elevated Hb values in blood measured in specific POC devices are not clear, but may include performance differences between the Hb measurement instruments, inherent variability in capillary blood samples, and differences in technique with distinct operators and protocols [Karakochuk 2019]. It is important to consider that these analytic instruments use different mechanisms to calculate Hb. For ADVIA-2120, blood samples are lysed within a microfluidic chamber by alkaline borate buffer and detected using cyanmethemoglobin chemistry to measure Hb levels [Malin 1989; Malin 1993; Bauer 2008]. For HemoCue 201+, blood samples are lysed within a cuvette by sodium deoxycholate and detected using the modified azidemethemoglobin chemistry to measure Hb levels [HemoCue 201]. For HemoCue 301, blood samples within a cuvette are directly measured for at Hb/HbO_2_ isobestic point to determine Hb levels [HemoCue 301]. It is possible that these differences in analytic method contribute to small discrepancies in Hb values from blood samples.

We also investigated the impact of BCT manufacturer on the measurement of Hb, comparing similar BCT products made by BD and Sarstedt. Using venous blood, mean Hb content was not significantly different between BD and Sarstedt BCTs, regardless of measurement using ADVIA-2120, HemoCue 201+, or HemoCue 301. The lack of a difference in Hb values with different BCTs may not be surprising given their similar construction. The 2 BCT types did have somewhat different anticoagulant additive, with BD using dipotassium ethylenediaminetetraacetic acid (K_2_EDTA) and Sarstedt using tripotassium ethylenediaminetetraacetic acid (K_3_EDTA), but these differences did not lead to different Hb values from venous blood. It was not possible to compare with capillary blood due to limited blood volumes. We are unaware of any previous study reporting comparing Hb values from different BCT manufacturers to compare our findings.

The results of this study indicate that both blood draw location and Hb measurement instruments can significantly influence the measurement of Hb. Although the overall difference is small, this influence can affect the assessment of anemia in target populations. For example, a recent study by Larson and colleagues showed that measuring Hb using capillary blood may overestimate the prevalence of anemia in the target population, leading to an incorrect classification of anemia severity in some of the subjects [Larson 2021]. Similarly, our study had 3 participants that were identified as mildly anemic due to Hb values below Hb threshold based on venous blood samples measured using an automatic hematology analyzer. Yet these same participants would not have been categorized as anemic if their capillary blood samples had been used instead, as they were higher than the Hb threshold. Also, 2 of the 3 participants would not have been categorized as anemic even based on venous blood if the samples had only been measured on HemoCue devices. Furthermore, the greater variance in Hb measurements observed with HemoCue devices compared to the ADVIA-2120 would only increase the likelihood of misidentifying the anemia status of these individuals when using POC instruments.

Although we attempted to minimize all extraneous variables, there were still some discrepancies that could have influenced the Hb values in this study. First, only 1 device for each HemoCue model was used for the study, so aberrant instrument performance issues cannot be ruled out completely. While both HemoCue devices always passed daily checks with external controls, the accepted performance range is rather wide and there is no mechanism for users to adjust the calibration of the HemoCue devices in the field [Karakochuk 2019]. Secondly, the collection of capillary blood was a less controlled procedure compared to venous blood collection, and sometimes there was enough volume to fill the capillary BCT to the recommended volume level. Lower volumes in the BCT results in a suboptimal anticoagulant to blood ratio, and the consequences of this condition on Hb measurement are not well defined. Finally, we used a different posture in our participants for collection of capillary (standing) versus venous (seated) blood. Posture is known to impact the level of Hb measured in blood samples, with standing favoring higher values [Lippi 2015; Chaudhary 2017]. It is unclear if this had an effect in our study, since participants were allowed to lean against a bench for support during the fingerstick and then move to a chair for phlebotomy with minimal time for changes in the distribution of blood volume. Subsequent studies on the analytic variables that affect Hb measurement may want to apply greater attention to these areas of concern. One non-standard procedure in this study was the choice of the thumb as a blood draw site. We selected the thumb due to several anecdotal reports that larger capillary blood volumes could be obtained compared to traditional fingerstick. Most published capillary sampling procedures either do not mention or specifically contraindicate the use of the thumb. Explanations for avoiding the thumb mostly claimed the pain would be higher to the blood donor, either due to the active use of the thumb or greater innervation compared to the fingers. However, we were unable to find any convincing references for these claims, and most participants in this study reported little to no pain with this approach. By using the thumb, we were able to routinely collect 1-2 ml of capillary blood from the participants. More detail on the use of the thumb for capillary blood sampling will be reported in a future publication.

The strength of this study is the control of the analytic variables outside of blood draw site and analytic device. Previous studies use multiple clinical locations, phlebotomists, processing times, and other factors because they had to contend with the challenges of large cohort and/or field assessments. Those concerns were minimized in this study so that the focus could be brought to the analytic variables of interest. The weakness of this study is also the use of healthy adult participants in a highly controlled environment, which does not reflect all the complexities of a large cohort and/or field assessment. Yet, it seems important to understand the base line variability inherent in different blood draw sites and analytic devices will inform the causes of variability in more complex studies. Another weakness is that this study was not designed with specific power to determine differences in analytic variables on Hb measurement, due to the ad hoc addition of Hb to a different set of analytes. It is likely that a larger study would refine the bias in blood draw sites and analytic devices, and may reveal important differences in other analytic variables.

## METHODS

### Study Overview

This study was conducted entirely on the campus of Children’s Hospital Oakland Research Institute (CHORI), now part of the University of California San Francisco. All clinical visits were scheduled within a 2-hour window (10:00AM–12:00PM). Participants completed a demographic questionnaire, anthropometric measurements, and non-fasting blood draw beginning with fingerstick(s) then immediately followed by phlebotomy. The blood samples were collected at our clinical site and then taken to our laboratory facility located less than 200 feet away with processing in 10-20 minutes on average and 60 minutes at maximum. Process variables for the blood draw and processing were minimized, including the use of the same clinical site, phlebotomist, blood draw procedures, and sample handling procedures. The CHORI Institutional Review Board approved this study (2019-055) in June 2019, and recruitment was conducted over 7 months.

### Study Participants

A diverse cohort of 60 healthy adults were recruited from the local community through flyers and word-of-mouth advertising. Inclusion criteria included being 18 years or older, generally healthy with no specified chronic illness or blood disorders. Exclusion criteria included not meeting inclusion criteria or being pregnant or lactating due to known physiological differences in nutrient homeostasis. Participants were asked to avoid consuming vitamin, mineral, and other supplements for 24 hours before their blood draw. Once at the clinical location, height and weight were measured using a Harpenden 602VR stadiometer and Scaletronix ST scale, which then were used to calculate body mass index (BMI) as mass (kg)/height (m) ^2^. Participants were also given a questionnaire asking for self-report of age, gender, and race/ethnicity use as qualitative markers of diversity. Race/ethnicity was merged to a single choice and also not used quantitatively since it is a social construct with little value in a scientific context. Participant demographics and questionnaire responses are provided (**Supplemental Table 1**).

### Clinical Procedures

#### Capillary blood collection

Participants were asked to stand up to maximize the effects of gravity on blood release from fingerprick, but lean against a workbench for comfort. Participant hands were warmed in a waterbath for 10-20 minutes and disinfected using alcohol wipes. Capillary blood was then collected by fingerstick using a BD high flow lancet (product number 366594, lot number V3V51E9) while participant remained standing. A fingerstick was applied to the non-dominant hand, first from the thumb and then 4 ^th^ finger if blood volume from thumb was insufficient. For explanation of the use of the thumb for capillary blood, see Discussion section. The phlebotomist gently pressed on thumb to start blood flow, but avoided any massaging, squeezing, or milking. The first drop of capillary blood was discarded, then remaining drops were pooled into either BD Microtainer 0.5 ml-size BCT containing K_2_EDTA (product number 365974, lot number 8311522) and Sarstedt Microvette 0.5 ml-size BCT containing K_3_EDTA (product number 20.1341.102, lot number 9482211). Capillary blood volume target was 0.5 ml or more, but was terminated short of target volume if blood from thumb and finger stopped before 0.5ml or participant requested to end the procedure. If more than 0.5ml was collected, multiple capillary BCTs would be used up to 2ml total volume. Once collected, tubes were inverted 4-6 times according to manufacturer’s instructions.

#### Venous blood collection

After capillary collection, the participant was then moved to a blood collection chair and seated position for phlebotomy [WHO 2010]. Venous blood was immediately collected by venipuncture in the arm of non-dominant hand, typically at the antecubital vein using a Sarstedt Safety Multifly Needle 21Gx3/4”-size with Multi-Adaptor (product number 85.1638.200, lot number 9050111), as this blood draw system accommodated both BD and Sarstedt BCT types. Venous blood was collected into BD Vacutainer 6 ml-size BCT containing K_2_EDTA (product number 368381, lot number 8187762) and Sarstedt Monovette 7.5 ml-size BCT containing K_2_EDTA (product number 01.1605.100, lot number 8031811).

### Laboratory Procedures

Blood samples were taken directly to the laboratory within 10-20 minutes on average, 60 minutes at maximum. Hb concentration was measured using an automated hematology analyzer ADVIA-2120 (Siemens Healthineers, instrument number IR14181915) and two point-of-care instruments HemoCue 201+ (HemoCue AB, serial number 0515012279) and HemoCue 301 (HemoCue AB, serial number 1246820215). The ADVIA-2120 was tested each run-day with Siemens 3-in-1 TESTpoint Hematology Controls according to the manufacturer’s instructions and were always within target range. The CV of the controls processed on 31 separate days were 0.81% for low standard, 2.72% for medium standard, and 1.21% for high standard. The HemoCue 201+ was tested each run-day with R&D Systems CBC-7 HemoCue kit (product number HC724, lot number R0819) with low, normal, and high Hb standards according to the manufacturer’s instructions and were always within target range. The HemoCue 301 was tested each run day with Eurotrol HemoCue 301 kit with level 1 (product number 188.001.002, lot number 92277), level 2 (product number 188.002.002, lot number 90778), and level 3 (product number 188.003.002, lot number 92279) Hb standards according to the manufacturer’s instructions and were always within target range. The ADVIA-2120 withdrew approximately 175µl of anticoagulated whole blood directly from the BCT through the manual open-tube sampler port. The ADVIA-2120 performed a complete blood count (CBC) analysis that included Hb measurement, and also flagged any blood sample with out-of-range hematological values. For the HemoCue 201+ and HemoCue 301 instruments, approximately 10µl of anticoagulated whole blood was manually pipetted into model-specific HemoCue cuvettes according to the manufacturer’s instructions and immediately analyzed. Replicate samples were processed for each whole blood sample (if volume allowed) and the average of the values was reported. Hb values exceeding 19 g/dL were flagged as improbable values and excluded from data summary.

### Statistics

The analysis of Hb was added ad hoc to an existing study on the differences in zinc concentration between capillary and venous blood samples, to be described in a later report. Thus, the participant number was not based on achieving specific power requirements for discriminating differences in Hb measurements from blood draw site or measurement device. Graphs and standard statistical testing were conducted using Prism 9 (GraphPad Software, Inc.). Interrelationships between variables were examined in a Pearson correlation matrix and Bland-Altman analysis. Outlier analysis was conducted using the GraphPad ROUT algorithm with Q=0.1% [Motulsky 2006]. Assessment of data normality utilized the D’Agostino-Pearson omnibus K2 normality test. Statistical significance was assigned at p<0.05.

## Data Availability

All data produced in the present study are available upon reasonable request to the authors.

## Abbreviations

ANOVA: analysis of variance test
BCT: blood collection tube
BD: Becton, Dickinson and Company
CBC: complete blood count
CHORI: Children’s Hospital Oakland Research Institute
CV: coefficient of variation
K_2_EDTA: dipotassium ethylenediaminetetraacetic acid
K_3_EDTA: tripotassium ethylenediaminetetraacetic acid
Hb: hemoglobin
LMIC: low and middle-income countries
NHANES: National Health and Nutrition Examination Surveys
POC: point-of-care
WHO: World Health Organization

## ACKNOWLEDGEMENTS

The authors thank Nyla Sepulveda for expert phlebotomy service and always making our participants feel comfortable during this study. The authors also thank Bonny Alvarenga and Timothy Jang for laboratory assistance, Dr. Ellen Fung for providing clinical space, Dr. Donnie Whitehead for use of a HemoCue 301 device, and Amy Letcher for donating Sarstedt tubes. Early work from this study was presented at the 2019 HEmoglobin MEasurement (HEME) Working Group Meeting, Arlington, VA, with many helpful comments from the committee members.

## FIGURE LEGENDS

**Supplemental Table 1.**
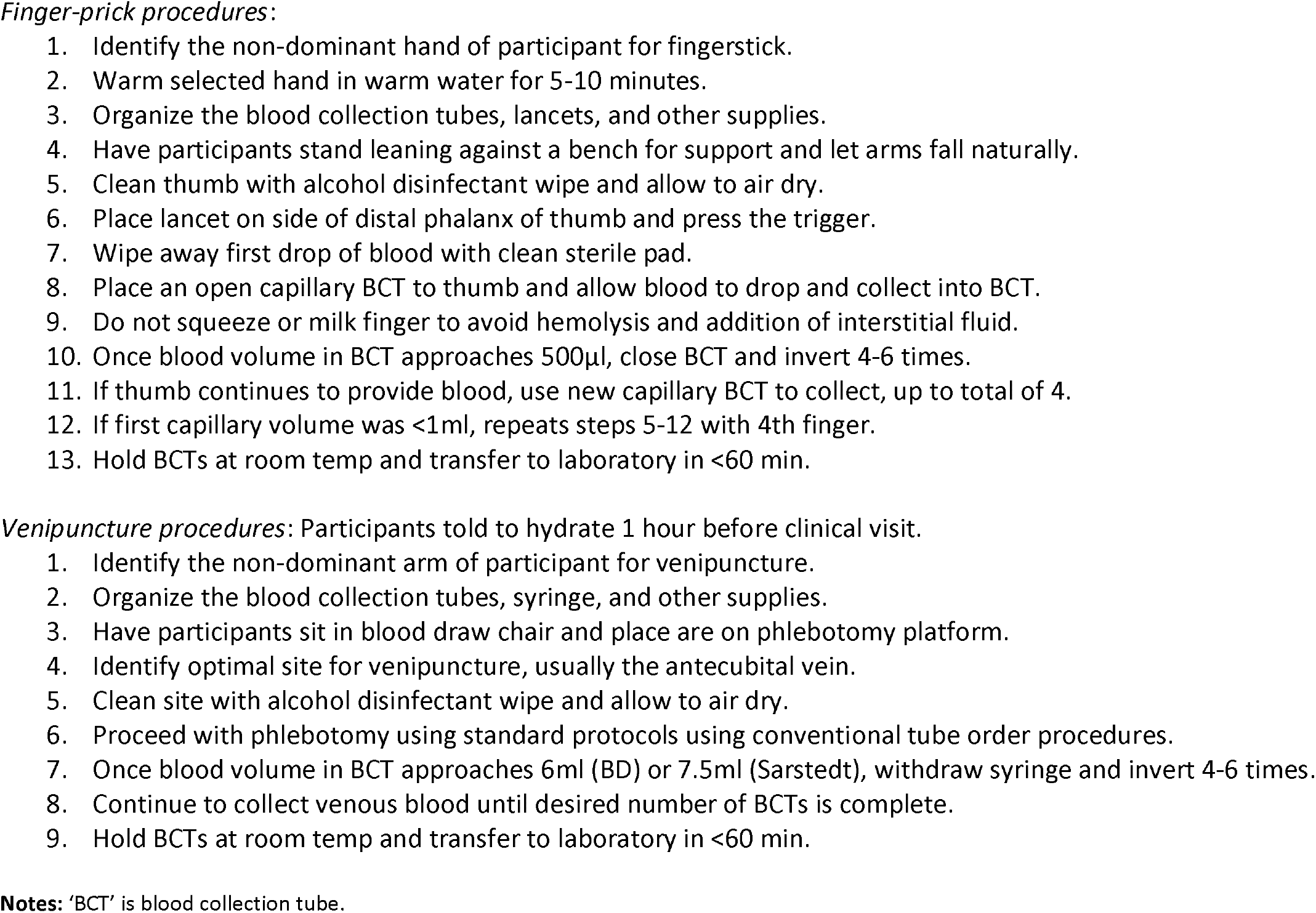
Procedure for venous and capillary blood draw.

**Supplemental Table 2.** Demographic information on study participants.

| participant # | age range | gender | race/ethnicity | supplement use | BMI |
| --- | --- | --- | --- | --- | --- |
| 1 | 40-49 | female | White | occasional | 19.0 |
| 2 | 40-49 | male | White | never | 41.0 |
| 3 | 50-59 | female | White | frequently | 19.6 |
| 4 | 50-59 | female | White | frequently | 33.6 |
| 5 | 30-39 | female | White | daily | 30.6 |
| 6 | 18-29 | female | Asian | rarely | 27.4 |
| 7 | 60-69 | male | White | occasional | 31.5 |
| 8 | 18-29 | female | Hispanic | occasional | 20.0 |
| 9 | 50-59 | male | AA/Black | daily | 32.7 |
| 10 | 50-59 | female | AA/Black | frequently | 19.3 |
| 11 | 30-39 | female | White | never | 21.2 |
| 12 | 40-49 | male | Hispanic | rarely | 21.4 |
| 13 | 60-69 | female | White | occasionally | 23.7 |
| 14 | 18-29 | male | AA/Black | never | 31.1 |
| 15 | 30-39 | male | Asian | never | 22.5 |
| 16 | 40-49 | female | White | rarely | 29.1 |
| 17 | 50-59 | male | White | occasionally | 23.5 |
| 18 | 18-29 | female | Mixed | never | 24.4 |
| 19 | 30-39 | male | Asian | never | 22.0 |
| 20 | 18-29 | male | Hispanic | daily | 29.3 |
| 21 | 18-29 | female | White | never | 27.3 |
| 22 | 60-69 | male | White | occasionally | 24.8 |
| 23 | 40-49 | female | Hispanic | never | 26.1 |
| 24 | 30-39 | male | White | never | 20.9 |
| 25 | 50-59 | female | AA/Black | rarely | 24.1 |
| 26 | 50-59 | female | Asian | daily | 22.2 |
| 27 | 18-29 | male | Mixed | frequently | 39.5 |
| 28 | 50-59 | male | Asian | occasionally | 25.1 |
| 29 | 40-49 | male | White | rarely | 25.3 |
| 30 | 30-39 | female | Asian | rarely | 29.3 |
| 31 | 30-39 | female | Asian | rarely | 21.6 |
| 32 | 60-69 | female | Mixed | daily | 44.4 |
| 33 | 50-59 | male | Asian | never | 30.4 |
| 34 | 60-69 | male | White | daily | 23.6 |
| 35 | 40-49 | female | Mixed | occasionally | 26.1 |
| 36 | 18-29 | male | Hispanic | frequently | 25.8 |
| 37 | 30-39 | male | AA/Black | never | 26.9 |
| 38 | 18-29 | female | Mixed | daily | 21.5 |
| 39 | 18-29 | male | Asian | never | 21.8 |
| 40 | 60-69 | male | White | occasionally | 25.0 |
| 41 | 18-29 | female | Asian | rarely | 33.1 |
| 42 | 18-29 | male | Mixed | rarely | 31.8 |
| 43 | 18-29 | female | White | never | 20.7 |
| 44 | 18-29 | female | Asian | daily | 17.1 |
| 45 | 18-29 | male | Hispanic | occasionally | 22.1 |
| 46 | 18-29 | female | Hispanic | rarely | 32.7 |
| 47 | 30-39 | male | Asian | never | 25.4 |
| 48 | 60-69 | female | White | occasionally | 34.3 |
| 49 | 18-29 | female | White | never | 21.0 |
| 50 | 30-39 | female | Asian | occasionally | 19.4 |
| 51 | 18-29 | male | White | occasionally | 25.3 |
| 52 | 40-49 | female | AA/Black | never | 23.4 |
| 53 | 40-49 | female | White | frequently | 27.4 |
| 54 | 40-49 | female | Middle Eastern | daily | 20.7 |
| 55 | 30-39 | male | Asian | occasionally | 25.5 |
| 56 | 18-29 | female | Asian | occasionally | 22.0 |
| 57 | 60-69 | male | White | daily | 34.3 |
| 58 | 18-29 | female | Asian | rarely | 22.1 |
| 59 | 60-69 | male | White | daily | 27.1 |
| 60 | 40-49 | female | White | daily | 29.2 |

**Notes:** Demographic and anthropomorphic data collected from study participants. ‘Mixed’ is identified as mixed race/ethnicity.

## Supplemental Figure Legends

**Supplemental Figure 1.**
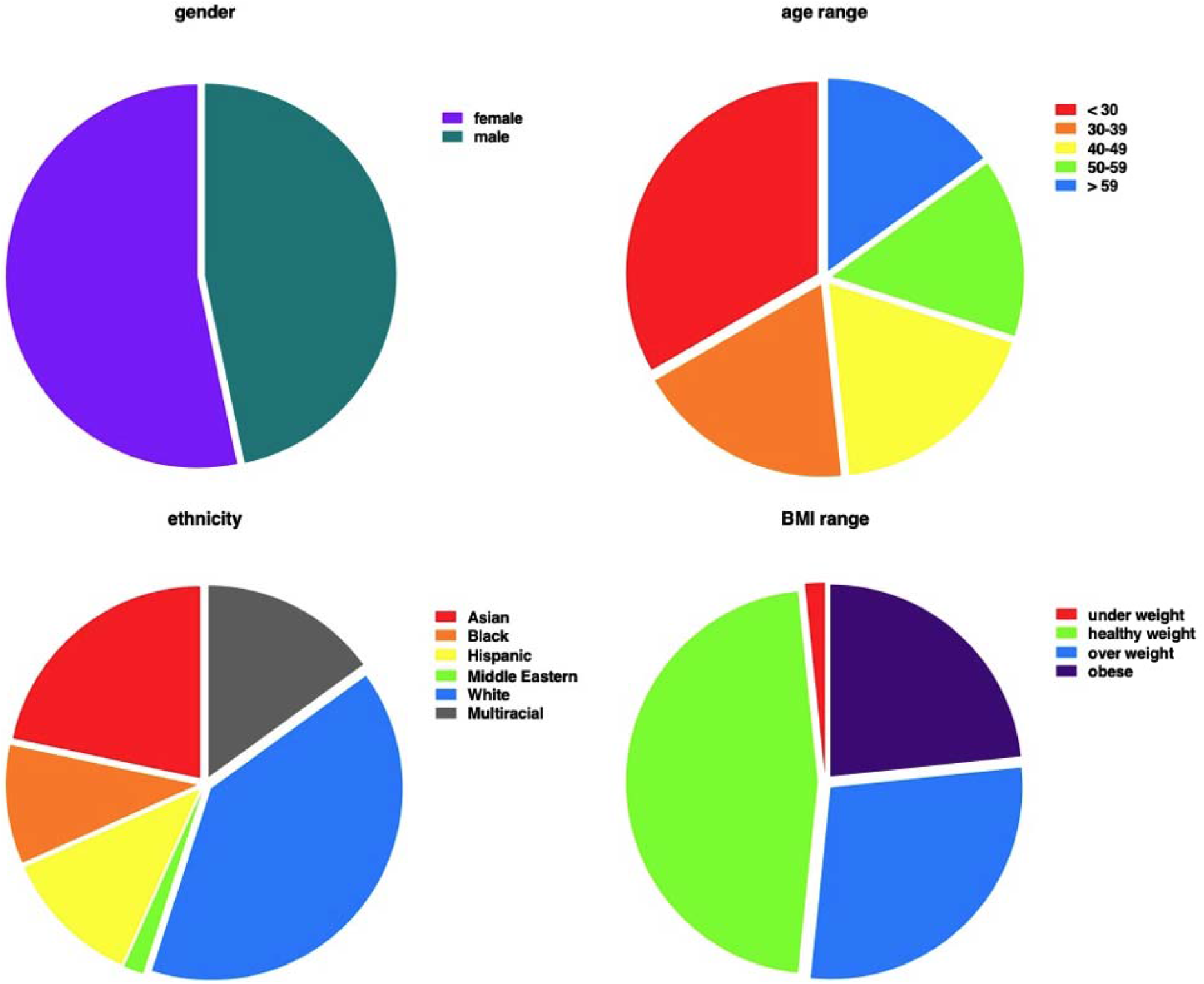
Summary of demographic information on study participants. Pie chart of gender, age-range, race/ethnicity, and body mass index (BMI) for participants. Gender, age, and ethnicity were provided directly by participants. BMI was calculated from height and weight measured on each participant.

**Supplemental Figure 2.**
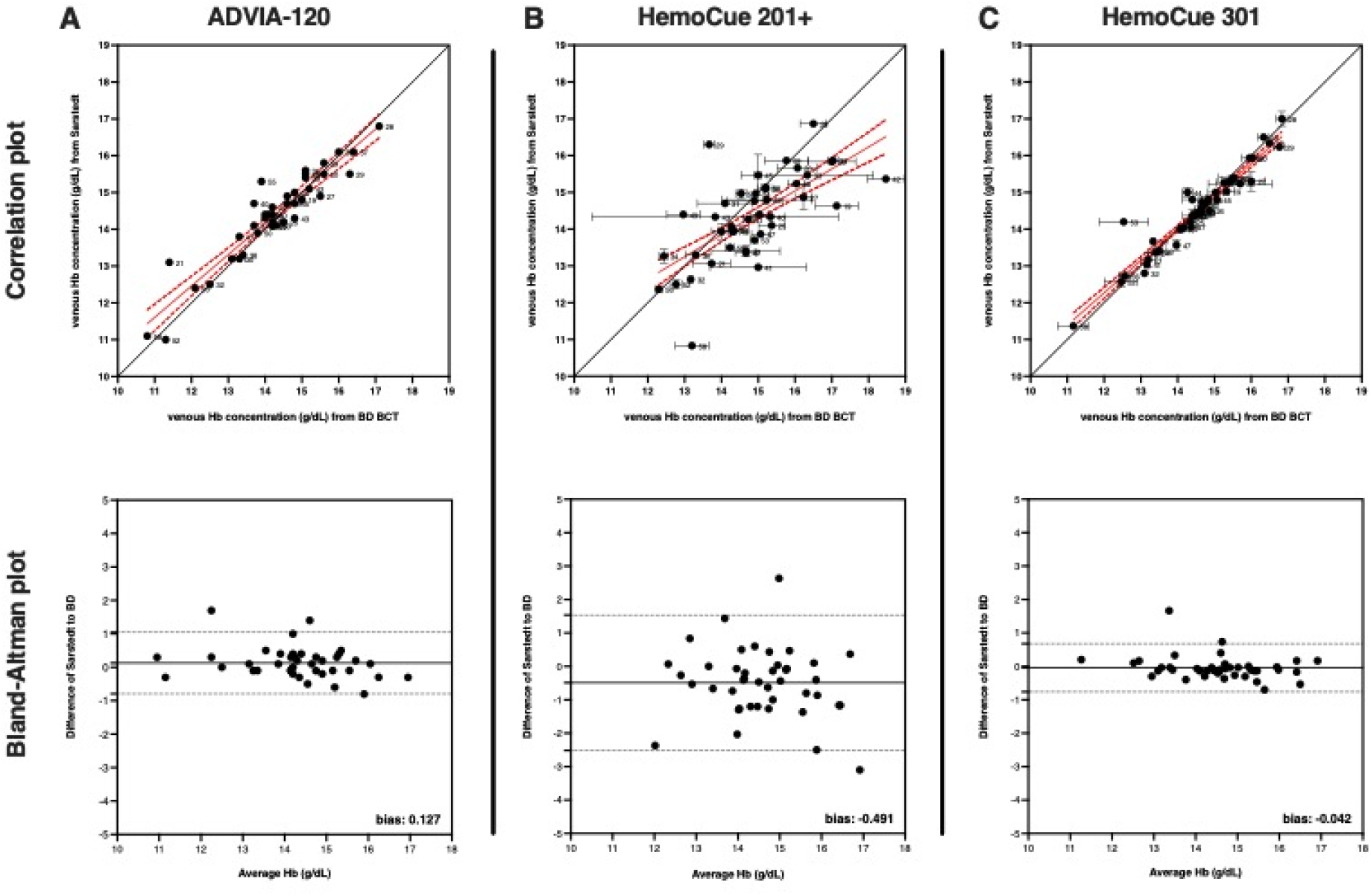
Hemoglobin concentration is not different in blood collected with blood collection tubes (BCTs) from different manufacturers. Correlation and Bland-Altman plots for Hb values determined in venous blood collected in BCTs made by BD or Sarstedt using (**A**) ADVIA-2120, (**B**) HemoCue 201+, and (**C**) HemoCue 301. The correlation plots show the capillary and venous Hb values (mean ± SEM) proximity to line of concordance (solid black line), as well as linear regression (solid red line) ± 95% confidence interval (red dotted line) for the group. Each circle represents a single participant, with participant ID number to the immediate right. The Bland-Altman plot shows a bias of (**A**) 0.127 ± 0.469 g/dL for Sarstedt over venous BD for ADVIA-2120, but a bias of (**B**) 0.491 ± 1.033 g/dL and (**C**) 0.042 ± 0.368 g/dL for HemoCue 201+ and HemoCue 301, respectively.

